# Uncovering the compounding effects of COVID-19 and racism on mental health disparities among biomedical PhD and MD students

**DOI:** 10.1101/2021.04.29.21251164

**Authors:** Allison Schad, Rebekah L. Layton, Debra Ragland, Jeanette Gowen Cook

## Abstract

The increasing visibility of mental health challenges for academic and graduate trainee populations has led to discussion of the role higher education institutions should play to address trainee mental health, particularly during the COVID-19 pandemic and ongoing racial injustice. To address the growing concern about training impacts on medical and biomedical doctoral trainee mental health, a cross-sectional study (n=957) was conducted using institutional annual survey data analyzed by type of training program, race/ethnicity, and survey year on measures of depression, anxiety, hazardous alcohol use, problems related to substance use, and suicidal ideation. Results indicated significant differences for rates of depression, anxiety, and suicidal ideation, with biomedical doctoral trainees showing greater incidence than medical doctoral trainees, and underrepresented minority trainees showing greater incidence than well-represented trainees. The concerningly high rates of depression, anxiety, and suicidal ideation among these trainee populations suggest that medical and biomedical doctoral training environments must be transformed in addition to expanding mental health support resources.

## Introduction

Graduate and professional training in the biomedical and health sciences provide a stressful environment for junior researchers and clinicians who are crucial to the future of our biomedical workforce. Yet, the majority of student mental health studies to date have focused on undergraduates, and when graduate students are included, disparate disciplines are combined. Considering that students in medical school and in biomedical graduate programs may be exposed to unique training experiences and stressors compared with other educational populations, we posited that they may also exhibit unique patterns in their mental health and well-being. In contrast to undergraduate, graduate, and professional students in many other disciplines, the requirements for in-person experiential learning for medical and biomedical trainees, either in clinical settings or in laboratories, precluded a transition to fully online instruction during the pandemic. Prior to COVID-19, reports indicated that medical students begin their medical education with lower rates of burnout and depression symptoms, and higher quality of life scores than similarly aged college graduates and the general population (Rosal et al, 1997; Brazeau et al, 2014). Yet, medical student mental health declines during medical school and is worse than the general population by graduation (Dyrbe 2006; Brazeau et al, 2014). Furthermore, the effects of the training experience on mental health may have been impacted by the 2020 COVID-19 pandemic. Unfortunately, there is a dearth of research on the mental health of biomedical PhD students. This study explores the mental health of medical students and biomedical doctoral trainees at the intersection of the COVID-19 pandemic and during continued racial injustice by comparing trainee self-reported mental health in 2020 as compared with 2019, corresponding with before versus during the widespread recognition of these dual crises in the public sphere.

In comparison to undergraduates and medical students, less is known about the mental health of biomedical PhD students. Nonetheless, the studies to date strongly indicate poor mental health among graduate students in general. One study found that graduate students at a Canadian Medical School have higher anxiety and depression than medical students (Toews et al, 1993), and an institutional graduate student survey found that approximately half of survey respondents reported emotional or stress-related problems (Hyun et al, 2006). Recent reports indicate elevated rates of anxiety and depression persist among graduate students at US and international universities (UC Berkley, 2014; Evans et al, 2018; Nagy et al, 2019; Levecque et al, 2017). The 2018 international study of PhD and Master’s degree-seeking students by Evans and colleagues found that found that 39% of respondents had indicators of moderate to severe depression, and 41% of respondents had moderate to severe anxiety scores (Evans et al, 2018). Stressful aspects of training environment, such as long work hours, pressures to produce, and unsupportive relationships with advisors may negatively impact PhD student mental health (Hazell et al, 2020). Additionally, financial concerns, uncertainty about future employment, and lack of clarity about university policies also contribute to PhD student stress, which may negatively impact mental health (Mackie & Bates, 2019). We wanted to understand mental health effects of the COVID-19 pandemic on medical students as well as on biomedical researchers in the lab, who may have potentially been disrupted collectively as a group across realms of scientific productivity, educational progression, and personal life. This disruption may have been more pronounced for biomedical researchers pursuing a PhD than some disciplines (e.g., humanities or social science PhD students) and differently than medical trainees (e.g., MD students), who could perhaps continue academic progress with some remote research or instruction without the same magnitude of disruption of academic progress.

Since the early 2000s, diagnosis of mental health disorders and demand for mental health counseling services on college campuses have increased (Benton et al, 2003; Oswalt et al, 2018; LeViness et al, 2019). This need was evidenced by a multi-campus study of combined undergraduate and graduate students, which found that 17.3% of students scored positive for depression, 9.8% scored positive for anxiety, and 6.3% reported suicidal ideation (Eisenberg, Hunt, & Speer, 2013). Of note, in some studies Asian, Black, Hispanic, and multi-racial undergraduate students scored higher for depression than white students (Eisenberg, Hunt, & Speer, 2013). Many theories have attempted to account for such racial and ethnic disparities – for instance, the effects of structural racism (Kendi, 2019) can impact symptoms, diagnosis, and treatment at many levels (Legha & Miranda, 2020). Examples of these multi-level impacts include the use of white populations for diagnosis without efforts to expand population norms (lack of scaling using white baseline as the norm); racial bias toward diagnoses of psychosis in black populations without any evidence of genetic predisposition; and documented systemic lack of access to care (for a review, see Conrad, 2020).

In addition to the highly stressful nature of the biomedical disciplinary training period for medical and graduate students, the 2020 COVID-19 pandemic resulted in globally increased symptoms of anxiety, depression, post-traumatic stress disorder, psychological stress, and stress, particularly among healthcare workers, those with pre-existing mental health conditions, females, college students, and individuals younger than forty-years-of-age (Xiong et al, 2020). The full consequences of this chronic stress on student mental health are not yet known.

Furthermore, the stress caused by the 2020 COVID-19 pandemic coincided with heightened responses and protests to the persistent racial injustice and violence in the United States. Large protests of lethal violence against marginalized people – notably killings of Black Americans by police and vigilantes – engulfed many US cities during a particularly contentious and racially-charged election season. Compounding systemic inequities and racial injustice, COVID-19 has disproportionally impacted communities of color in the United States (Li, 2020; Hooper et al, 2020; CDC, 2020). These unique combinations in 2020 may have exacerbated mental health disparities among trainees, particularly for Black and Indigenous People of Color (BIPOC) who are clinicians and scientists. We investigate comparisons between well-represented (WR) and under-represented (URM) groups in medical and biomedical PhD students.

During the COVID-19 pandemic, medical and doctoral trainee experiences were impacted in multiple domains such as limited social interaction; increased threat of illness; uncertainty of future employment; and drastic changes to research, training, and clinical schedules. These and other factors could have caused disproportionate effects on the mental health of medical and biomedical trainees, in particular. Trainees of color, especially people of color identifying as Latinx, Black, and Indigenous peoples, may also be affected by heightened concerns about individual, community, and family health and safety due to the disproportionate impact of COVID-19 on communities of color paired with systemic racial inequity and the threat of racial violence. The aim of the current study is to assess changes in the mental health status of medical and doctoral trainees during the COVID-19 pandemic and a time of reckoning of persistent racial injustice.

## Methods

The School of Medicine at a large public university in the Southeastern United States conducts annual voluntary surveys of both medical students and biomedical PhD students. Comparisons were made using two comparable data collection timepoints with identical items on the survey, both administered at a similar time of year, distributed using the same mechanisms (e.g., same internal listservs), and both open for similar periods of time. In order to better understand trainee mental health, we compared survey data from 2019 and 2020.

The annual survey was administered to MD and biomedical PhD students (n=431, Fall 2020; n=526, Fall 2019). Each survey was open for four weeks between September and October, approximately one year apart so as to be as comparable as possible. The study was reviewed and approved by the Institutional Review Board (#18-0112). Mental health data was analyzed by type of training program (MD vs. PhD), race/ethnicity (well-represented vs. underrepresented), and year (2019 vs. 2020).

## Measures

We measured depression, anxiety, hazardous alcohol use, and problems related to substance use with widely utilized and validated instruments: the Patient Health Questionnaire (PHQ-9), Generalized Anxiety Disorder Assessment (GAD-7), Alcohol Use Disorder Identification Test (AUDIT), and the Drug Abuse Screening Test (DAST). Three additional questions were asked to assess student suicidal ideation (ever, while enrolled, and past 12 months). Demographic data including race/ethnicity were also collected. In some cases, partial survey data was recorded (n=957 total responses; of those, n=740 completed survey). Partial surveys were used, however, only fully completed measures were included in the analysis (a blocked survey design enabled a data cleaning check that ensured participants completed each section/measure they were working on before closing the survey).

Categories for mental health measures were calculated using validated measures such that increasing numbers indicate higher category levels of severity. For each of the four measures, category zero indicated a lack of presentation of symptoms or no problematic substance use, as applicable. Other numbers (1-3 or 1-4) indicated increasing population-normed-levels of severity for each variable (see **Appendices 1-7 for Supplemental Figures and Tables** with full categorical distributions annotated).

Depression categories were calculated (PHQ-9; Kroenke, Spitzer, & Williams, 2001) with summed values of responses sorted into categories ranging from 0-4, using definitions: 0) 1-4: Minimal or No Depression, 1) 5-9: Mild Depression, 2) 10-14: Moderate Depression, 3) 15-19: Moderately Severe Depression, 4) 20-27: Severe Depression. Anxiety categories were calculated (GAD-7; Spitzer et al 2006) with summed values of responses sorted into the following categories, ranging from 0-3: 0) 0-4: Minimal Anxiety, 1) 5-9: Mild Anxiety, 2) 10-14: Moderate Anxiety, 3) 15-21: Severe Anxiety. Hazardous Alcohol use was assessed (AUDIT; World Health Organization, 2001) with summed scores sorted into the following categories: 0) 0-7: Low Risk, 1) 8-15: Hazardous, 2) 16-19: Harmful, 3) 20+: Possible Dependence. Problems with drug use was assessed as use of prescribed or over the counter medication in excess of their directions and any non-medical use of drugs (e.g., problems with drug use as defined by DAST-10; Skinner, 1982; Yudko, Lozhinka, & Fouts, 2007) and was summed into the following categories: 0) 0: No problems, 1) 1-2: Low Problems, 2) 3-5: Moderate Problems, 3) 6-8: Substantial Problems, 4) 9-10: Severe Problems. Suicidal ideation was assessed using the following question: “Have you ever thought about ending your life?” and respondents indicated *Yes* or *No* for each of the following: *Ever, While Enrolled*, and *In the Last 12 Months*. For all analyses (see **Figures** below), all measures were recoded as a bivariate (0/1) variable with one indicating symptoms or problems with drug use and zero the lack thereof, respectively.

### Participants

Respondents included MD and biomedical PhD students (n=431, Fall 2020; n=526, Fall 2019) of approximately 800 MD students and 600 PhD students who were invited to participate. Respondents identified as the following: 66% White, 14% Asian, or 2% Other (coded as Well Represented, WR); and 7% African American, 7% Latinx, 2% Middle Eastern, 1% American Indian/Alaskan Native, and <1% Pacific Islander (collectively coded as Underrepresented, UR if indicating any of these identities). The majority respondents identified as female (67%), followed by male (32%), and <1% other (genderqueer, gender nonconforming, gender non-binary, and trans). Institutionally the overall MD and PhD student populations are more female than male, so these distributions are not unexpected. Respondents identified as straight/heterosexual (83%), bisexual (7%), gay/lesbian (5%), queer (2%), pansexual (2%), asexual (1%), or other (1%). Ages ranged from 18-40+, in the following age categories: with the majority ages 21-25 years (59%) and ages 26-30 (35%), with the remainder ages 31-35 (5%), 18-20 years (<1%), and 35-40 years (<1%).

## Results

### Effects by Year

Due to different patterns emerging for training type and racial/ethnic identity, limiting evaluation by year obscured these differences. To better elucidate the effects of each variable on mental health outcomes using a single model, a Logistic Regression was conducted. Additionally, post-hoc Chi-Squared tests are conducted within each sub-population. Finally, percentages across sub-populations are compared to better operationalize changes over time and by variable.

### Logistic Regression Model

Each mental health outcome variable was split into clinically meaningful bivariate categories for depression, anxiety, problems with drug use, and hazardous alcohol use such that the baseline category (none or fewest symptoms, as defined by each scale) was coded as zero (0), and any symptoms as defined by more or worse symptoms than the baseline category were coded as one (1). A bivariate logistic regression model (see **Appendix 1 – Supplemental Table 1**) was used to assess the impact of our primary factors of interest across the biomedical trainee populations to maintain a large sample size and sufficient power to compare well-represented and under-represented groups pre and during COVID-19. Year was coded into a practically meaningful bivariate category, with pre-COVID-19 as zero (2019, 0) and during the COVID-19 pandemic and racial unrest as one (2020, 1). Finally, well-represented and under-represented status were coded into a practically meaningful bivariate category, with endorsement of any UR category being coded as one (1), whereas all others were coded as well-represented using the value of zero (0).

There were significant changes by year, as well as differences between WR and UR trainee mental health outcomes for depression and anxiety. In the combined population there was a decrease in depression and anxiety (likely driven by the MD trainee improvements; see **Discussion**). In general, UR trainees exhibited significantly higher rates of depression and anxiety at about 1.5 times the rate of their WR peers across a combined MD plus biomedical trainee sample. Finally, UR trainees were approximately 2 times as likely to say they had thought about ending their life in the past 12 months than their WR peers, and about 1.8 times more likely while enrolled; such high rates are *extremely* concerning.

### Chi-Squared Post-Hoc Analyses

Follow-up analyses were further evaluated using a post hoc 2×2 Chi-Squared between each population (MD versus PhD trainees), as well as within each population (MD-only or PhD-only) to compare trends by racial identity (WR vs. UR trainees) and Year (2019 vs. 2020; see **Appendix 2 – Supplemental Table 2**). Mirroring effects on the Logistic Regression model, Depression, Anxiety, and Suicidal Ideation *While enrolled*, and *In the Past 12 months*, evidenced significant patterns, as summarized below.

#### Depression and Anxiety

Surprisingly, for MDs, there was a significant *decrease* in depression and anxiety over time (*p*s<.001), whereas for PhDs we observed no change between years. However, PhD UR trainees differed from their WR peers in that they were significantly more likely to be depressed *(p*<.03); UR trainees did not differ from their WR peers on measures of anxiety, problems with drug use, or hazardous alcohol use. MD UR trainees did not differ for depression or anxiety compared with WR peers.

#### Problems with Drug Use and Hazardous Alcohol Use

No significant differences of note emerged between populations or within populations on these measures. Reported problems with substance use and hazardous alcohol use were comparatively low in contrast to depression and anxiety, which were more pervasive.

#### Suicidal Ideations

MD showed trends toward improvement, whereas PhDs exhibited no change between years. There was a significant *increase* in MD UR trainee suicidal ideation *while enrolled* (*p*=.03), and PhD UR trainee suicidal ideation that indicated they had thought about ending their life in the *Past 12 months* (*p*=.01).

### Medical school versus biomedical doctoral training

Prior to 2020, both MD and PHD trainees suffered from depression (MD: 46%, PhD: 65%) and anxiety (MD: 47%, PhD: 67%) at high rates (see **Appendix 3 - Supplemental Figure 1a** and **Appendix 4 - Supplemental Table 3**), as defined by no symptoms compared with any symptomatic categories (e.g., 0 versus combined categories 1, 2, 3, 4; see **Methods** for categorical definitions). PhD trainee mental health in 2020 remained very poor (depression MD: 26%, PhD: 64%; anxiety MD: 32%, PhD: 61%), whereas surprisingly, MD trainee mental health improved during 2020. Suicidal ideation in the past 12 months among PhD trainees (compared with MD trainees) was markedly higher, both before (MD: 11%, PhD: 16%) and during 2020 (MD: 6% PhD: 19%). We used chi-squared tests to assess significant differences between MD and PhDs for depression (PHQ-9), anxiety (GAD), problematic drug use (DAST), and hazardous alcohol use (AUDIT) using nominal outcome variables (see **Appendix 4 – Supplemental Table 3**). We identified significantly higher rates of depression and anxiety for PhD trainees compared with MD trainees, as well as higher rates of suicidal ideation *while enrolled* and *over the last 12 months* (see **Table 1, Appendix 3 - Supplemental Figure 1b**, and **Appendix 4 - Supplemental Table 3**). No significant differences emerged for problems with drug use, or hazardous alcohol use by training type, racial/ethnic identity, or year; furthermore, problems with drug use rates were comparatively low in contrast to depression and anxiety.

### Well-represented versus underrepresented trainees

Mental health results for well-represented (WR) versus underrepresented (UR) across a combined pool of biomedical research trainees and medical trainees were also compared (see **Table 1**). UR trainees experienced higher rates of depression and anxiety compared to their WR peers, in both 2019 (depression, WR: 51%, UR: 60%; anxiety, WR: 53%, UR: 60%), and 2020 (depression, WR: 35%, UR: 48%; anxiety, WR: 38%, UR: 51%; see **Table 1**; see also **Appendix 5 - Supplemental Figures 2a and Appendix 6 - Supplemental Table 4**). Suicidal ideation was also worse for underrepresented trainees as compared with well-represented peers, specifically *while enrolled* and *last 12 months* (2019 WR-UR 15% vs. 23% and 11% vs. 19%; 2020 WR 10% vs. 18%, 12% vs.17%, respectively). In summary, UR outcomes in general were worse for depression and anxiety (see **Figures 1 & 2**; see also **Appendix 5 - Supplemental Figures 2a & 2b**, and **Appendix 6 - Supplemental Table 4**) as well as experiencing more suicidal ideation *while enrolled* and *over the last 12 months* (see **Figure 3** and **Appendix 7 – Supplemental Tables 3a & 3b**).

**Figure 1.**
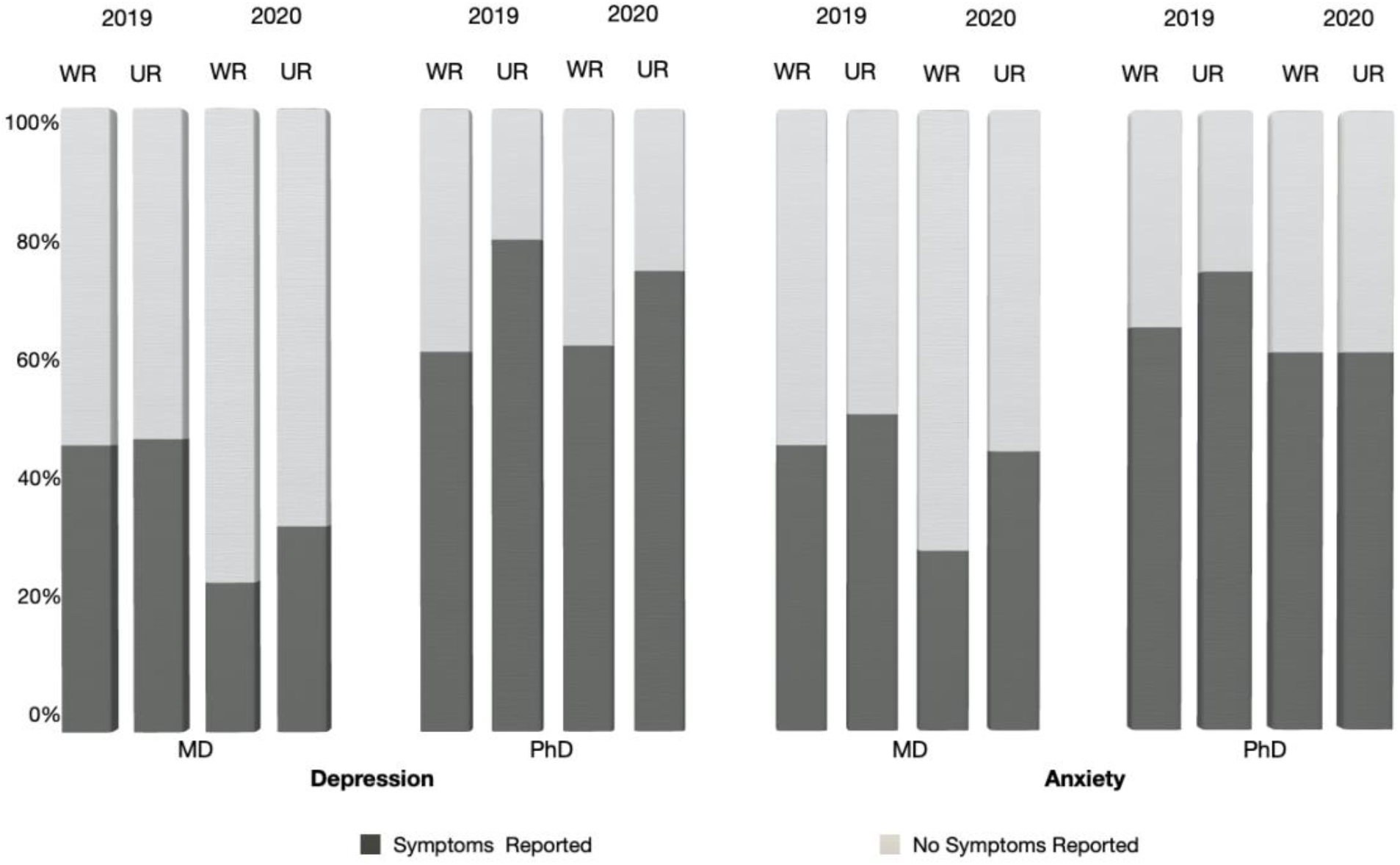
Depression and Anxiety for WR vs. UR MD and PHD in 2019 and 2020. All percentages were calculated out of total valid responses (n=622 MD, n=309 PhD; see **Appendix 7 - Supplemental Tables 3a & 3b** for *n*s and percentages). After coding measures into meaningful clinical categories ranging from 0-4 (Depression) or 0-3 (Anxiety) respectively, each categorical scale was re-coded into a bivariate 0/1 indicating the absence or presence of the respective symptoms for Depression and Anxiety (see **Measures**).

**Figure 2.**
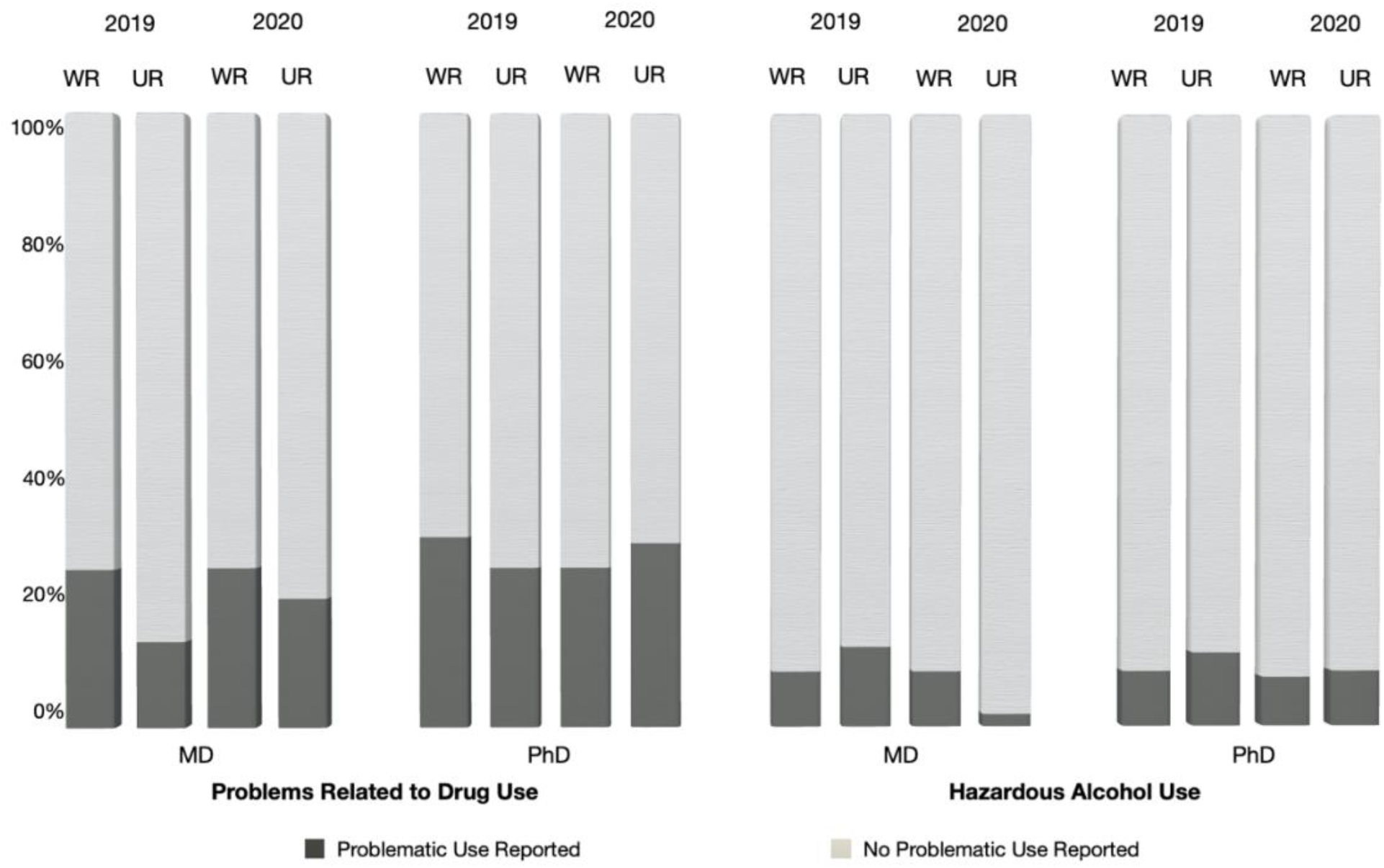
Problems with Drug Use and Hazardous Alcohol Use for WR vs. UR MD and PHD in 2019 and 2020. All percentages were calculated out of total valid responses (n=622 MD, n=309 PhD; see **Appendix 7 - *Supplemental Tables 3a & 3b*** for *n*s and percentages). After coding measures into meaningful clinical categories ranging from 0-4 (Problems with Drug Use) or 0-3 (Hazardous Alcohol Use) respectively, each categorical scale was re-coded into a bivariate 0/1 indicating the absence or presence of the respective symptoms for Problems with Drug Use or Hazardous Alcohol Use (see **Measures**).

**Figure 3.**
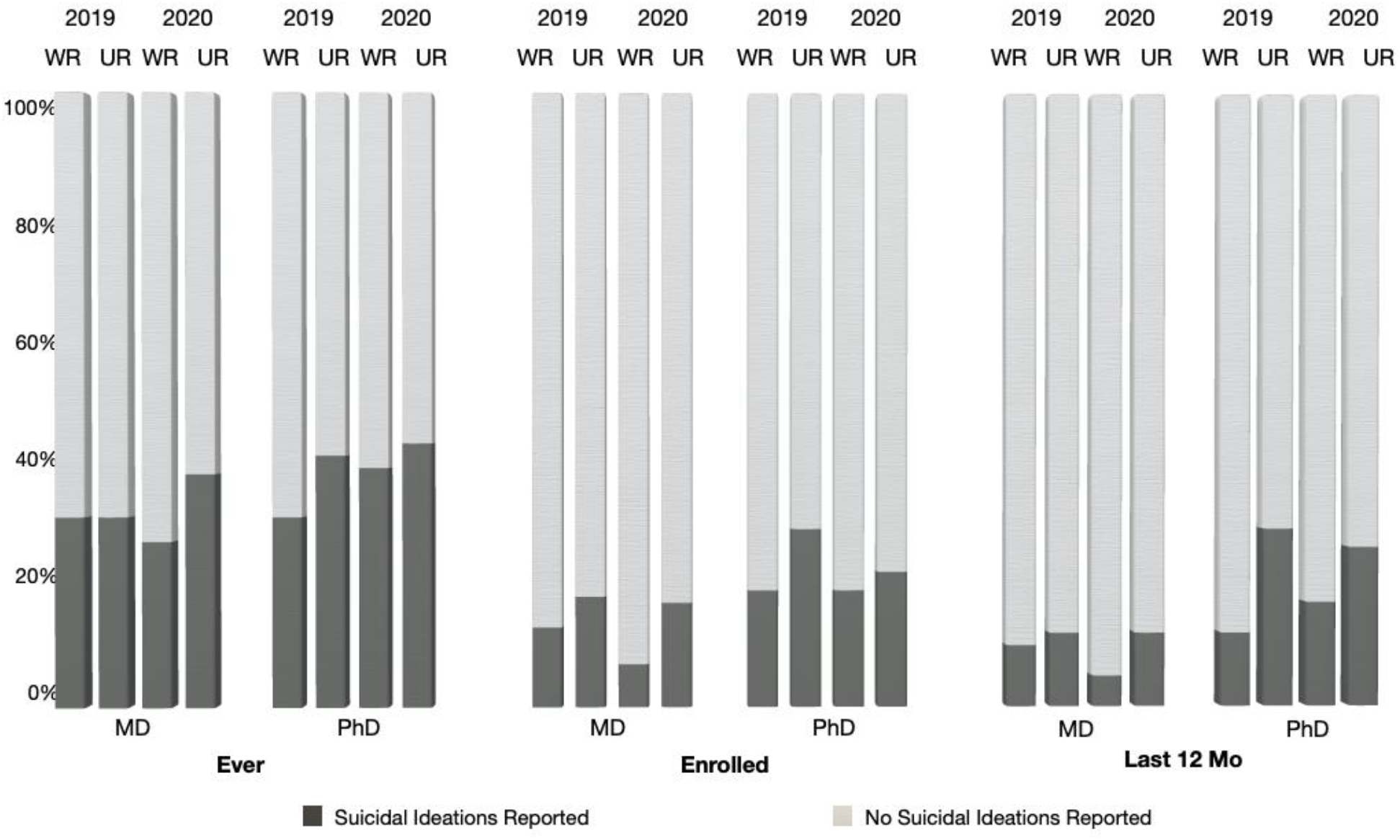
Suicidal ideation for WR vs. UR MD and PHD in 2019 and 2020. All percentages were calculated out of total valid responses. Response options included “Yes” (indicating suicidal ideations) or “No” (indicating no suicidal ideations) for each of the three categories (see **Methods** for exact question wording; see **Appendix 7 - Supplemental Tables 3a & 3b**).

**Figure 4.**
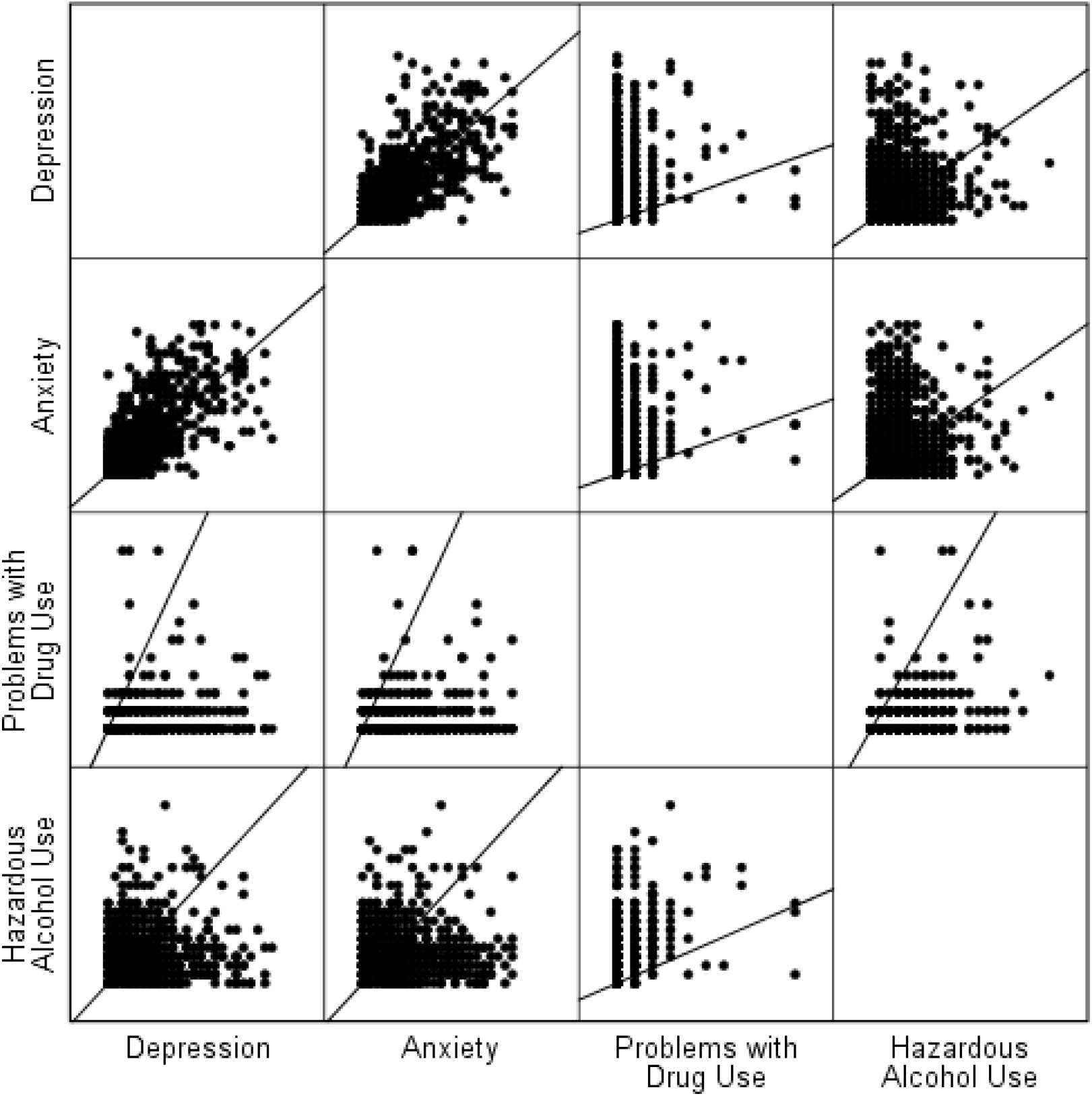
Associations between Depression, Anxiety, Problems with Drug Use and Hazardous Alcohol Use. Scatterplots of the relationship between variables of interest (Depression, Anxiety, Problems with Drug Use and Hazardous Alcohol Use) displayed include graphical representations in a matrix format.

### Associations

A robust positive association was evident between depression and anxiety (*r*=.69, *p*<.001), not surprisingly as these conditions are often comorbid. Both depression (*r*=.14) and anxiety (*r*=.33) were associated with problems with drug use (*p*s<.001). Neither depression (r=−.01, p=.74) nor anxiety (r=−.01, p=.77) were associated with hazardous alcohol use. Since this was only correlational in nature, a casual direction cannot be determined; nonetheless, these associations may indicate a connection between problems with drug use either as a precursor to or an effect of experiencing mental health symptoms.

## Discussion

Overall, MD trainee mental health improved significantly by year from 2019 to 2020 on measures of both depression and anxiety, whereas PhDs showed no significant change by year. UR trainees were worse off than their WR peers in measures of depression, anxiety, and suicidal ideation - particularly reporting more suicidal ideation than their peers both as MDs *while enrolled* and as PhDs in the *last 12 months*.

### Medical trainee trends

Despite the unique challenges between 2019 and 2020, many metrics of MD student mental health improved. In mid-March of 2020, medical students were pulled from clinical settings and from typical coursework due to the pandemic. Usual academic and clinical responsibilities were replaced with a four-week online course, “Medical Management of COVID-19,” focused on COVID-19 (e.g., wellness, self-care, medical management, PPE, virus spread, epidemiology, and other modules; UNC Health & UNC School of Medicine, 2020). It is possible that the pause in traditional medical student training and the new course focused on timely health topics had mitigating effects on stress-induced mental health erosion. We also speculate that being in a medical training program with obvious direct impact during a time when medical professionals were publicly celebrated had a positive effect on MD student mental health. Furthermore, medical students were able to continue some form of their training remotely, maintain some social contact and peer support networks through academic training, and avoid graduation delays. Additionally, while medical trainees pay for their training, which could contribute to financial stress, the financial demands of education did not change between 2019 and 2020. This consistency of circumstances combined with a pre-determined graduation date that was not delayed by the pandemic, expectations of financial income post-graduation, and a near-guarantee of eventual employment in medicine compared to peers in other fields could also have mitigated the downward trend in MD student mental health.

### Biomedical research trends

In contrast to MD students, biomedical PhD students exhibited alarmingly high rates of depression and anxiety that dwarfed the rates in the MD student population, which was higher than the general population. When considering recent suicidal ideation in particular, PhD student mental health worsened between 2019 and 2020. Biomedical PhD student training was severely curtailed in the 2020 spring months, and students returned to labs in June 2020 with strict occupancy limits that reduced the supportive social interactions and camaraderie among peers that typify most research groups. The loss of progress on dissertation projects given that graduation times are open-ended was frustrating and may have exacerbated mental health issues. In addition, the concurrent economic recession may have depressed optimism about job opportunities for PhD students, who pursue a much wider range of careers after graduation compared to MD students. Exacerbating this was the dismissal of science by politicians and in the media, popular sentiments against following public health policies based on scientific recommendations, and public degradation of trust in science and scientists, all of which may have contributed to a worsened experience for biomedical scientists.

### Medical education vs. biomedical doctoral training

During the COVID pandemic, MD and PhD trainees experienced the impact on their lives quite differently. There were stark differences in the structure of the training programs and in how much the pandemic restrictions disrupted training for the two groups. For PhDs whose research was not directly COVID-19 related, it may have been harder to find meaning and purpose in their formerly satisfying research on cancer, heart disease, etc. Anecdotally some biomedical PhD trainees sought opportunities to volunteer their molecular biology skills in diagnostic COVID-19 labs, indicating a desire to contribute to the immediate crisis. In contrast, MD trainees may have found it easier to connect their training with the real-world crisis of COVID-19. Moreover, when labs were shutting down, medical education shifted to COVID-19 prevention topics directly related to the pandemic. Additionally, whereas medical trainees will soon become medical doctors in a society where this role is even more respected than before the pandemic, and often seen as heroic, PhDs did not experience a similar level of adulation for their efforts. We do speculate that as the successful vaccine effort takes center stage, basic research scientists may find themselves celebrated more than in previous years. Future directions of education and mental health research should delve into the aspects of medical education versus biomedical PhD training that drive different mental health outcomes both during a crisis and in normal times.

### Underrepresented trainee trends

Mental health metrics for depression, anxiety and suicidal ideation were worse for students from under-represented groups compared to students from well-represented groups irrespective of MD or PhD program and worsened even more between 2019 and 2020. We hypothesize that the COVID-19 pandemic and ongoing racial violence contributed to these differences, although future research is needed to isolate the factors causing the observed effects. Academic culture is based on cultural norms that systematically exclude minority groups and create additional mental health challenges for people of color (e.g., Persons Excluded due to Ethnicity or Race; PEER; Asai, 2020). Overall, the incidence of suicidal thoughts for UR trainees increased proportionately both *while enrolled* and *in the Past 12 months* compared with WR trainees, and particularly for PhD UR trainees in the *Past 12 months*. This effect may be partially explained by the added stressors of systemic racism in academic, medical, and American culture.

### Associations

Our findings that depression was correlated with both anxiety and with problematic use of drugs was consistent with previous work which demonstrates common comorbidity of depression and anxiety, as well as the comorbidity of drug use disorders with mental illness (e.g., NIDA, 2010); hence, problematic drug use as either a contributing factor to (or coping strategy to manage) depression may pose a plausible explanation, but should be interpreted with caution. Furthermore, the lack of association in our sample between hazardous alcohol use and mental illness was surprising given the common associations found between these variables in previous work (e.g., Smith & Randall, 2012; McHugh & Weiss, 2019).

## Conclusions

The mental health crisis in medical education is widely recognized, whereas the mental health crisis in graduate biomedical education has only recently received considerable attention from the laboratory research community. Resources have been mobilized to evaluate and respond to the needs of medical students that indeed still merit additional attention given high rates of depression, anxiety, and suicidality. These data suggest a need for swift action to similarly address the very urgent mental health needs of biomedical graduate trainees exhibited both before and during the COVID-19 pandemic, especially for UR PhD trainees. Furthermore, the data presented here suggests increases in depression, anxiety, and suicidality for UR medical and biomedical research trainees that have been exacerbated during the ongoing COVID-19 and racial inequity pandemics. Future studies should explore additional disparities between UR and WR trainees in addition to those highly concerning trends currently identified.

### Recommendations

It’s crucial, now more than ever, to provide mental health support both on-campus and remotely to ensure that students have access to the mental health services they need. On-campus mental health resources should reflect the racial and ethnic diversity of the student body. Mental health resources, communities, and support groups are starting to emerge online to help fill this need (e.g., <u>PhDBalance</u>, <u>TAE Consortium</u>, <u>RVoice</u>). While providing more resources is an important step, providing comprehensive mental health care to all of those in need may be untenable to continue in an uncontrolled scaling up to fully meet the increasing demand for mental health services seen on university and college campuses nationally (Seppälä et al, 2020). Therefore, programmatic preventative health measures should also be implemented and investigated at the graduate level, particularly regarding the learning environment. Further, due to inequitable access to these resources and systemic issues that negatively impact mental health outcomes, rather than simply providing services and programming, changes should be made to identify and address factors that drive the mental health crisis, particularly for BIPOC and biomedical PhD trainees given higher indicators of distress. Prevention, wellness resources, anti-racism, and resiliency training and assessment of impact are crucial to reduce the acute need for mental health support. More importantly, it is imperative that leaders in higher education use evidence-based research to identify and reduce causes of mental health problems and not only treat symptoms when they emerge. Some potential causes to explore include toxic work environments, systemic racism, and unhealthy cultural and academic norms. Higher education leaders should actively reform the academic culture to model, encourage, and sustain student, faculty, and staff wellbeing. Furthermore, evidence-based research should focus on the mental health of faculty and staff in addition to that of students, since faculty and staff who struggle with their own mental health may be poorly situated for effective teaching and student support.

### Limitations

First and foremost, response bias is always a consideration in cross-sectional self-report research. Relatedly, these were not matched controls, hence it is possible that sampling distributions may have differed by chance. Furthermore, it’s possible that respondents differed based on how important mental health is to them, potentially skewing the sample; hence we cannot definitely evaluate the respondent sample as representative of the full population. We achieved a response rate commensurate with voluntary survey data, suggesting a typical level of participation (250/800 MDs – 125/600 PhDs). We also had a high percentage of women respondents, who experience higher rates of depression and anxiety compared to male respondents; however, greater female response is not atypical of our graduate medical and biomedical trainee population, which includes more enrolled women than men. Even given the limitations of our data, the unexpectedly high rate of suicidal ideation is concerning. Among respondents alone, nearly 40 trainees (16 MDs and 21 PhDs) reported recent suicidal ideation in 2020.

We recognize that other social identity groups beyond race and ethnicity (Rothman, Gunturu, Korenis, 2020) may also experience inequitable impacts of COVID-19 and similar structural biases may negatively impact mental health for other reasons; for instance, gender identity and sexual orientation (Salerno et al, 2020), international and undocumented status (Chen et al, 2020), or disability (Goggin & Ellis, 2020; Gray et al, 2020). Future directions should include examination of the intersectionality of these and other identity groups, and systemic barriers that each may encounter differentially.

Furthermore, additional factors not accounted for herein may have also exacerbated the mental health status of trainees. For instance, while data collection was completed prior to the 2020 election, political tensions were building throughout the summer and fall of 2020, which may have also provided contributing factors impacting the well-being of trainees. Future research should examine the impact of political cycles and governmental policies on trainee populations (for instance the impact of xenophobic, nationalist, ableist, sexist, and homophobic legislation impacting international trainees, people of color, disabled people, women, and LGBTQ+ people in the United States), including election cycles, economic trends, and other federal policies.

## Data Availability

Data is shared in accordance with IRB approval in de-identified format and can be accessed via an Open Science Framework project.

https://osf.io/8bd7v/?view_only=5c9401974a2545e2a3516ce1a728f986

## Appendix 1.

**Supplemental Table 1.** **Chi-Squared by Training Type, Racial/Ethnic Identity, and Year**

| Training Type (MD vs. PhD) |  |  |  |  |
| --- | --- | --- | --- | --- |
| Category | Phi | ChiSq | P-val | Trend Description |
| **Depression (PHQ)\*\*\*** | **-0.27** | **60.81** | **<.001** | **PhD worse** |
| **Anxiety (GAD)\*\*\*** | **-0.24** | **44.31** | **<.001** | **PhD worse** |
| Problems with Drug Use (DAST) | -0.05 | 1.93 | 0.17 | No difference between MD and PhD |
| Hazardous Alcohol Use (AUDIT) | 0.01 | 0.04 | 0.84 |  |
| Suicidal Ideation - Ever | -0.06 | 3.11 | 0.08 |  |
| **Suicidal Ideation - While Enrolled\*\*\*** | **-0.13** | **13.56** | **<.001** | **PhD worse** |
| **Suicidal Ideation - Last 12 Mo\*\*\*** | **-0.13** | **13.24** | **<.001** | **PhD worse** |
| Racial/Ethnic Identity (WR vs. UR) |  |  |  |  |
| Category | Phi | ChiSq | P-val | Trend Description |
| **Depression (PHQ)\*\*** | **0.09** | **6.04** | **0.01** | **UR Worse** |
| **Anxiety (GAD)\*\*** | **0.08** | **4.78** | **0.03** | **UR Worse** |
| Problems with Drug Use (DAST) | -0.05 | 1.59 | 0.21 | No difference by race/ethnicity |
| Hazardous Alcohol Use (AUDIT) | 0.01 | 0.01 | 0.98 |  |
| Suicidal Ideations - Ever | 0.05 | 2.35 | 0.13 |  |
| **Suicidal Ideation - While Enrolled\*\*** | **0.09** | **6.42** | **0.01** | **UR Worse** |
| **Suicidal Ideation - Last 12 Mo\*\*\*** | **0.1** | **8.64** | **<0.01** | **UR Worse** |
| Year (2019 vs. 2020) |  |  |  |  |
| Category | Phi | ChiSq | P-val | Trend Description |
| **Depression (PHQ)\*\*\*** | **-0.16** | **21.23** | **<.001** | **Interaction** |
| **Anxiety (GAD)\*\*\*** | **-0.14** | **15.86** | **<.001** | **Interaction** |
| Problems with Drug Use (DAST) | -0.01 | 0.01 | 0.94 | No difference by year |
| Hazardous Alcohol Use (AUDIT) | -0.04 | 1.14 | 0.29 |  |
| Suicidal Ideation - Ever | 0.001 | 0.01 | 0.98 |  |
| *Suicidal Ideation - While Enrolled†* | -0.07 | 3.75 | 0.05 | *Interaction* |
| Suicidal Ideation - Last 12 Mo | -0.04 | 1.4 | 0.24 | No difference by year |

## Appendix 2.

**Supplemental Table 2.** **Logistic Regression Results**

Appendix 2. Supplemental Table 2. Logistic Regression Results
| Mental Health Category | Factor | P-Value Interval | Odds Ratio | 95% Confidence Interval |  |
| --- | --- | --- | --- | --- | --- |
|  |  |  |  | Lower Bound | Upper Bound |
| Depression (PHQ) | Year*** | <.001 | .52 | **0.39** | **0.69** |
|  | UR* | .02 | 1.55 | 1.08 | 2.23 |
| Anxiety (GAD) | Year*** | <.001 | .57 | **0.43** | **0.75** |
|  | UR* | .04 | 1.47 | 1.02 | 2.12 |
| Hazardous Alcohol Use (AUDIT) | Year | .29 | .75 | 0.45 | 1.27 |
|  | UR | <.99 | 1.00 | 0.53 | 1.89 |
| Problems with Drug Use (DAST) | Year | .91 | .98 | 0.71 | 1.36 |
|  | UR | .21 | .76 | 0.49 | 1.17 |

| Suicidal ideation | Factor | P-Value Interval | Odds Ratio | 95% Confidence Interval |  |
| --- | --- | --- | --- | --- | --- |
|  |  |  |  | Lower Bound | Upper Bound |
| Ever | Year | .95 | 1.01 | 0.75 | 1.36 |
|  | UR | .13 | 1.34 | 0.92 | 1.94 |
| While enrolled | Year | .06 | .68 | 0.45 | 1.02 |
|  | UR* | **.01** | **1.78** | **1.12** | **2.81** |
| Past 12 months | Year | .27 | .78 | 0.50 | 1.22 |
|  | UR* | **.01** | **2.05** | **1.25** | **3.35** |

## Appendix 3. Supplemental Figure 1a & 1b

**Supplemental Figure 1a.**
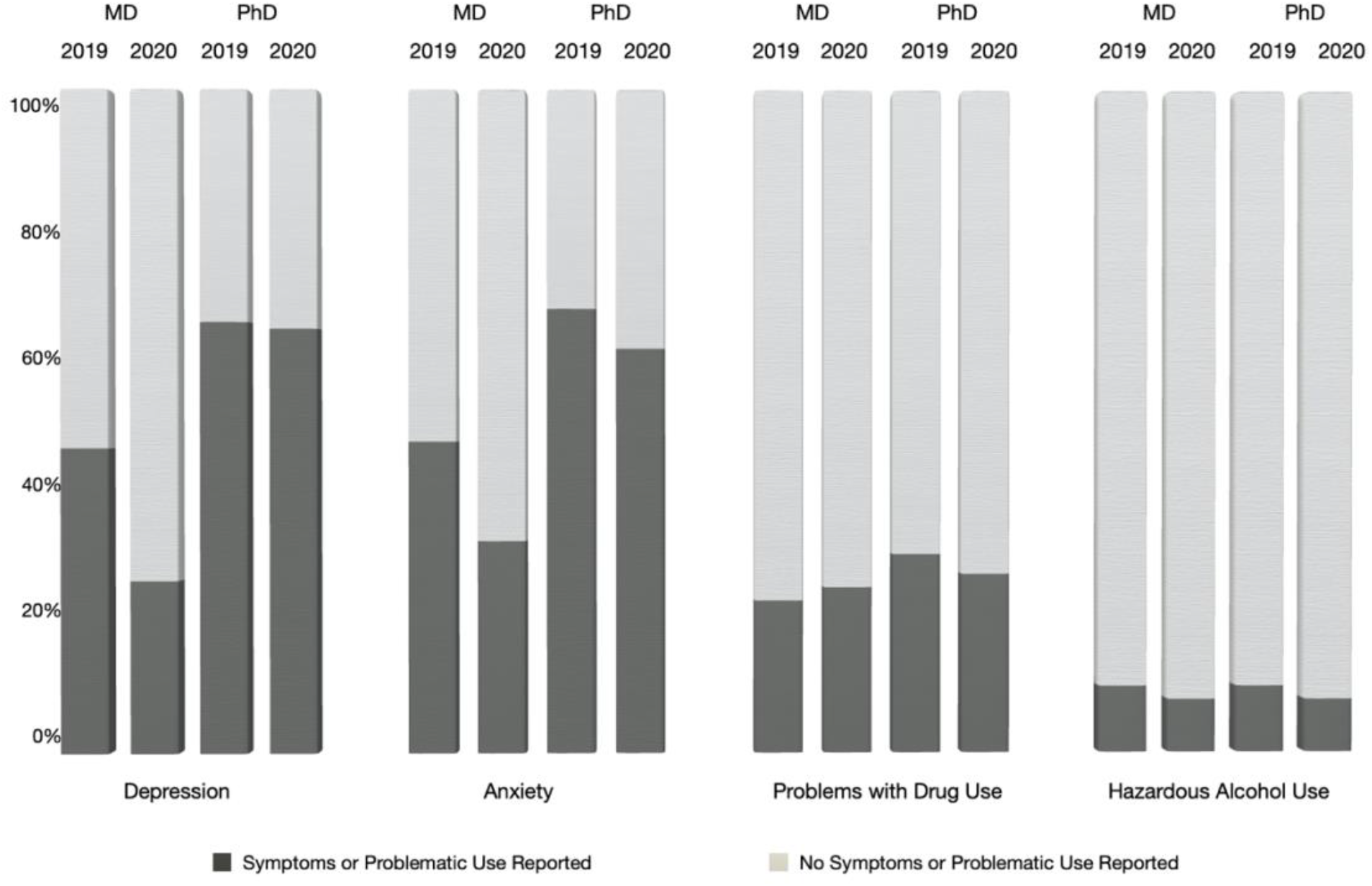
Depression, Anxiety, and Problems with Drug Use, and Hazardous Alcohol Use for MD vs. PHD 2019 and 2020. All percentages were calculated out of total valid responses (n=622 MD, n=309 PhD). After coding measures into meaningful clinical categories ranging from 0-4 (Depression, Problems with Drug Use) or 0-3 (Anxiety, Hazardous Alcohol Use) respectively, each categorical scale was re-coded into a bivariate 0/1 indicating the absence or presence of the respective symptoms for Depression, Anxiety, Problems with Drug Use, and Hazardous Alcohol Use (see **Measures**).

**Supplemental Figure 1b.**
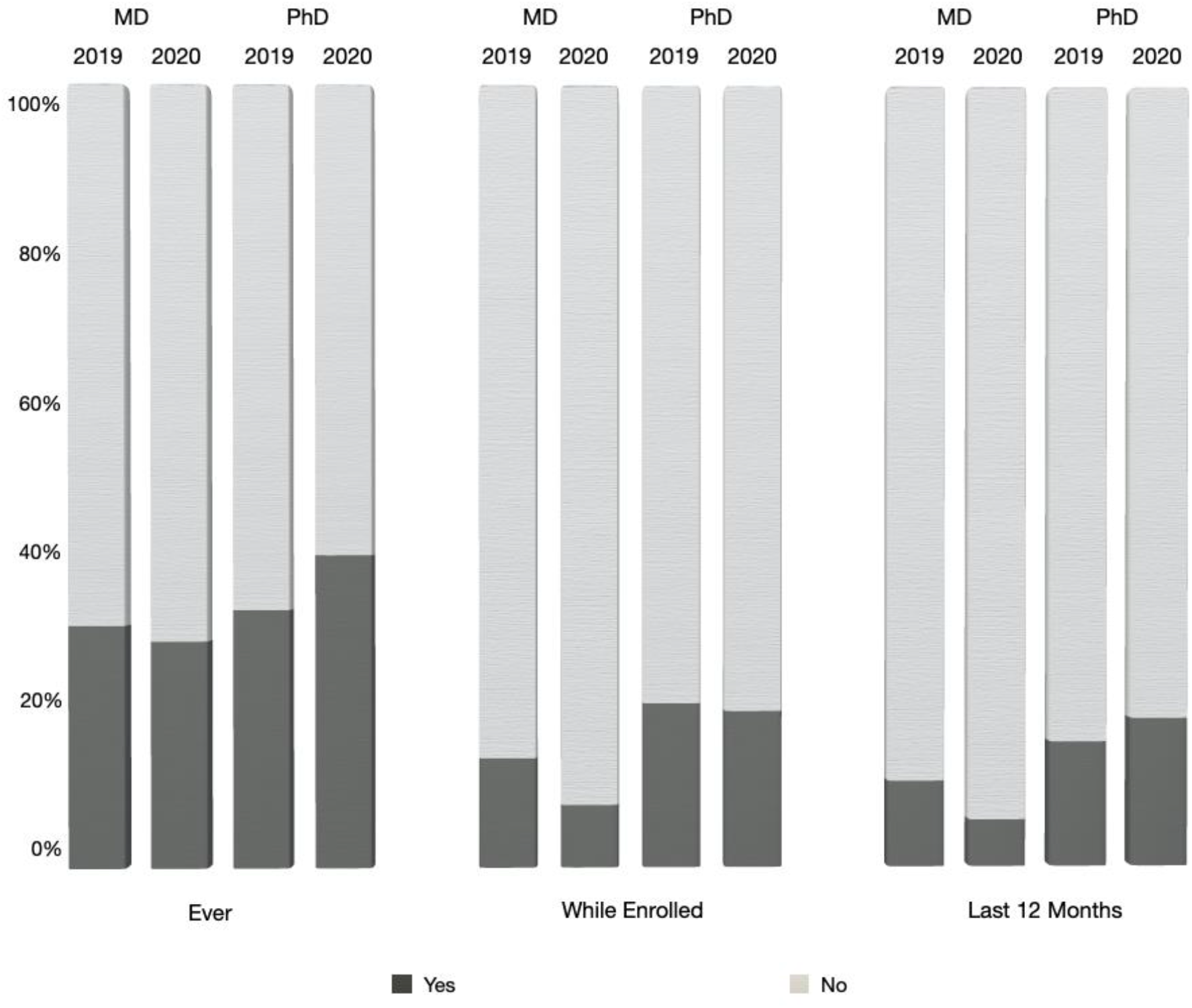
Suicidal ideation for MD vs. PHD 2019 and 2020. All percentages were calculated out of total valid responses. Response options included “Yes” (indicating suicidal ideations) or “No” (indicating no suicidal ideations) for each of the three categories (see **Methods** for exact question wording).

## Appendix 4.

**Supplemental Table 3.**
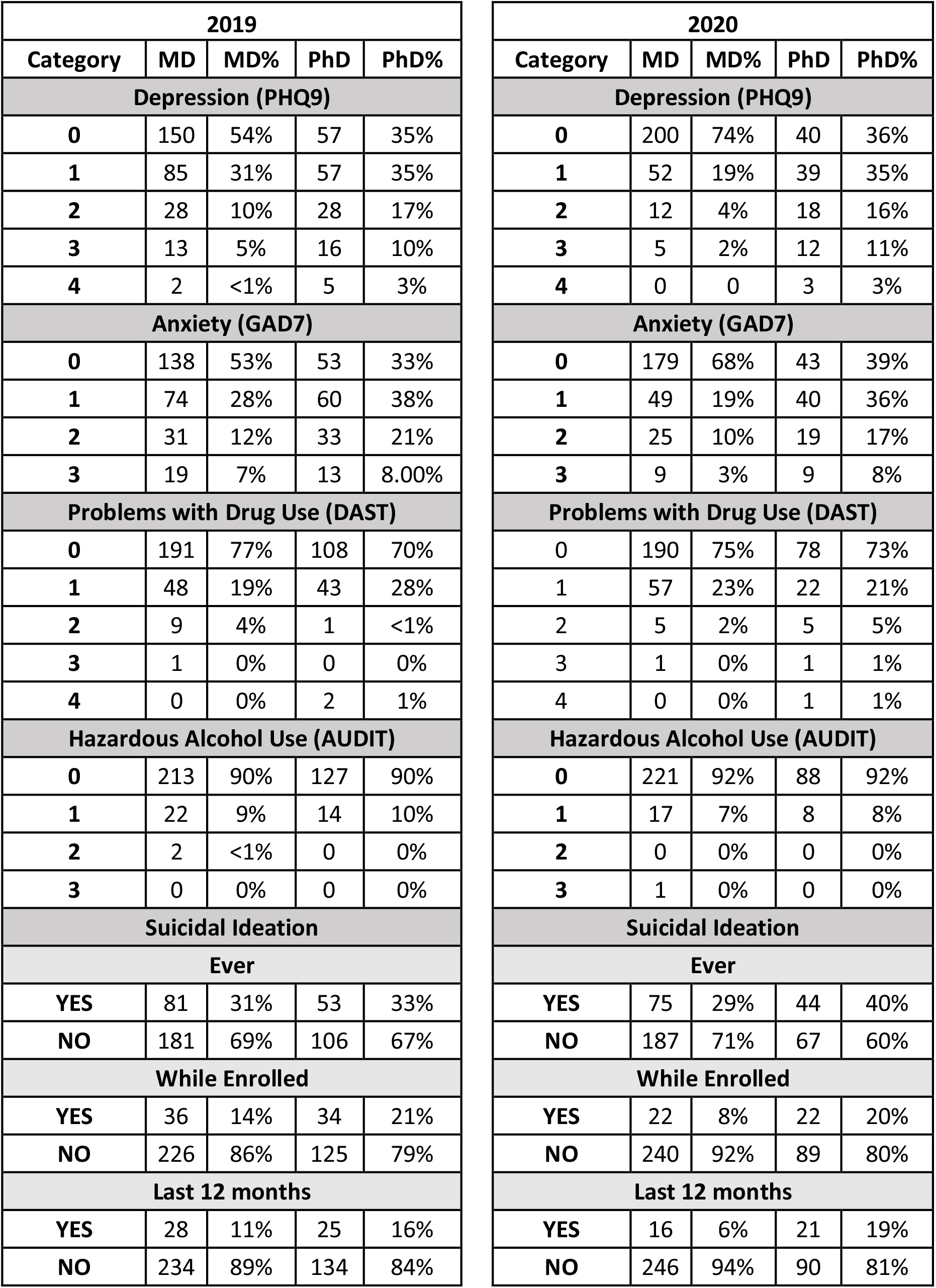
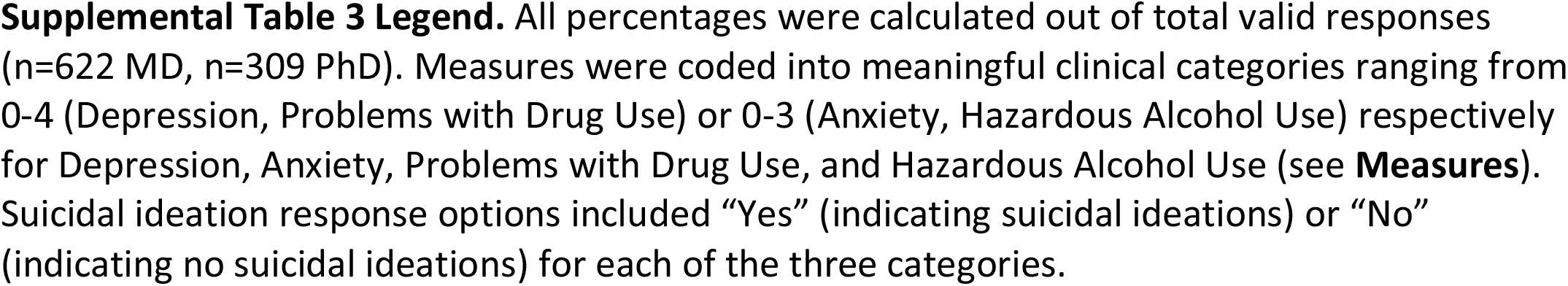
**Depression, Anxiety, Problems with Drug Use, Hazardous Alcohol Use, and Suicidal Ideation for MD vs. PhD Trainees**

## Appendix 5. Supplemental Figure 2a & 2b

**Supplemental Figure 2a.**
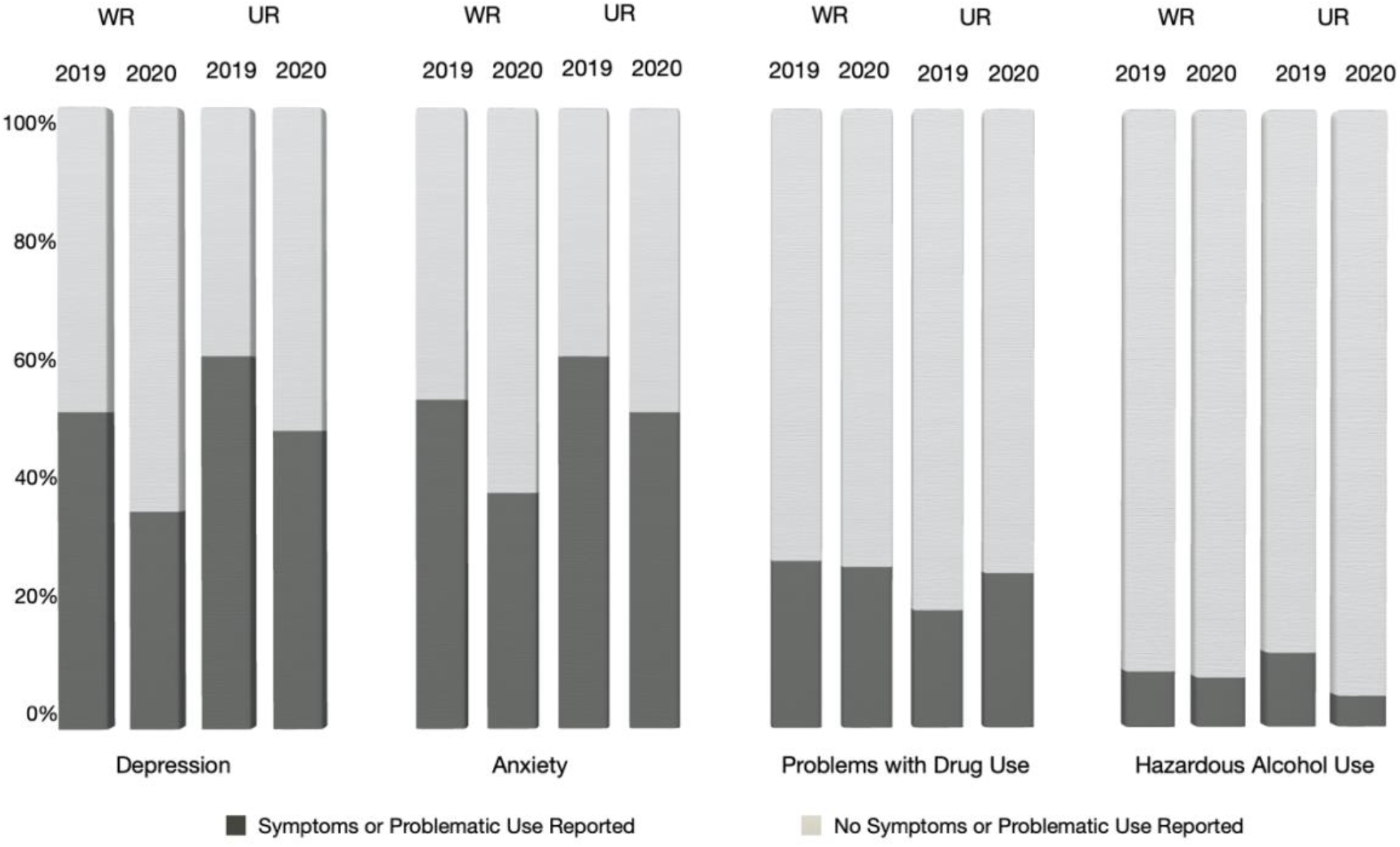
Depression, Anxiety, Problems with Drug Use, and Hazardous Alcohol Use for WR vs. UR 2019 and 2020. All percentages were calculated out of total valid responses (n=531 MD WR, n=91 MD UR; n=252 PhD WR, 57 PhD UR). After coding measures into meaningful clinical categories ranging from 0-4 (Depression, Problems with Drug Use) or 0-3 (Anxiety, Hazardous Alcohol Use) respectively, each categorical scale was re-coded into a bivariate 0/1 indicating the absence or presence of the respective symptoms or problematic substance use for Depression, Anxiety, Problems with Drug Use, and Hazardous Alcohol Use (see **Measures**). UR trainees were coded as such if they indicated racial or ethnic categories underrepresented in science (e.g., African American/Black, Hispanic/Latinx; see **Methods** for details), WR included any trainees who did not indicate a marginalized social identity.

**Supplemental Figure 2b.**
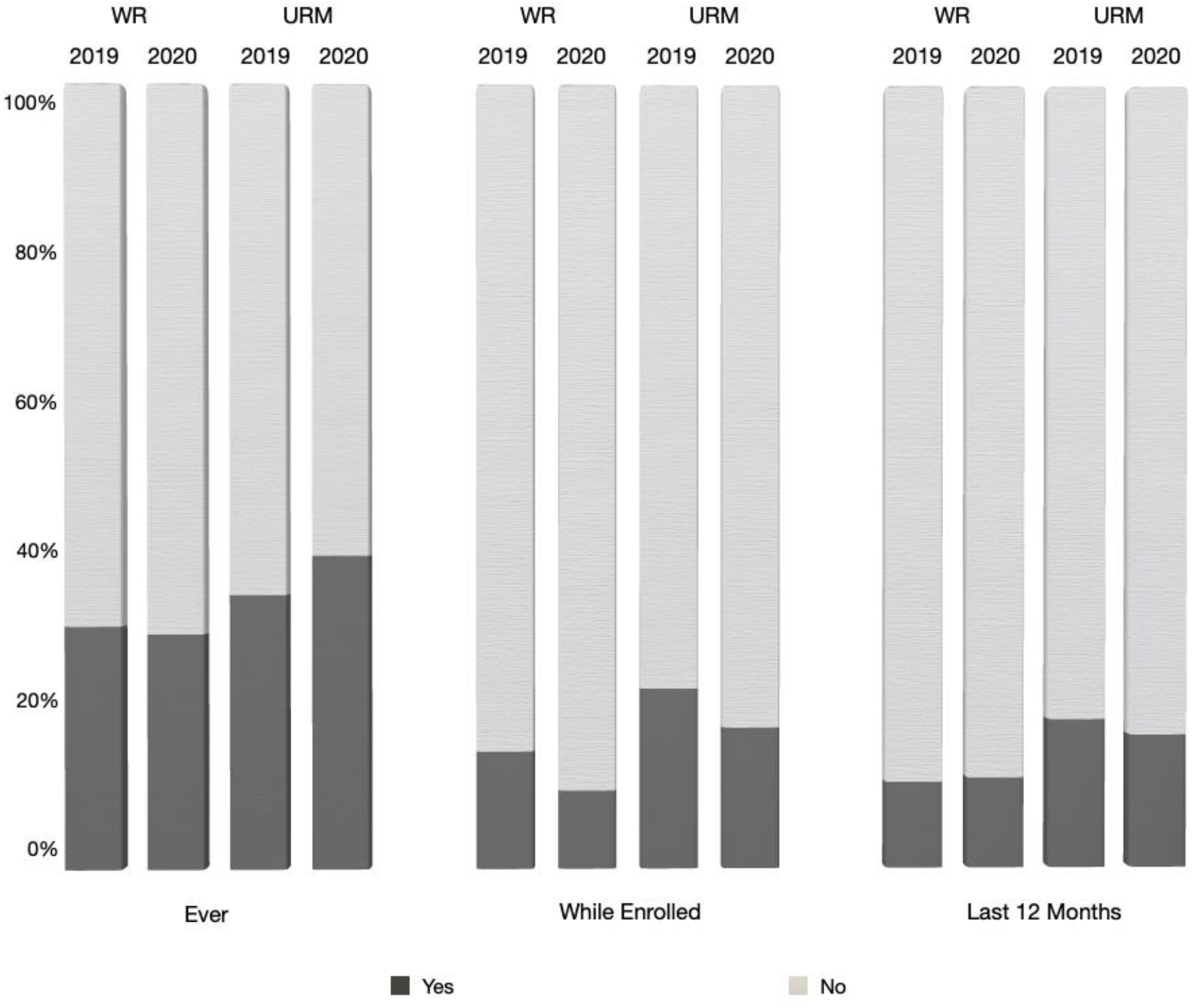
Suicidal ideation for WR vs. UR 2019 and 2020. All percentages were calculated out of total valid responses. Response options included “Yes” (indicating suicidal ideations) or “No” (indicating no suicidal ideations) for each of the three categories. UR trainees were coded as such if they indicated racial or ethnic identities underrepresented in science (e.g., African American; see **Methods** for details), WR included any trainees who did not indicate a marginalized social identity.

## Appendix 6.

**Supplemental Table 4.**
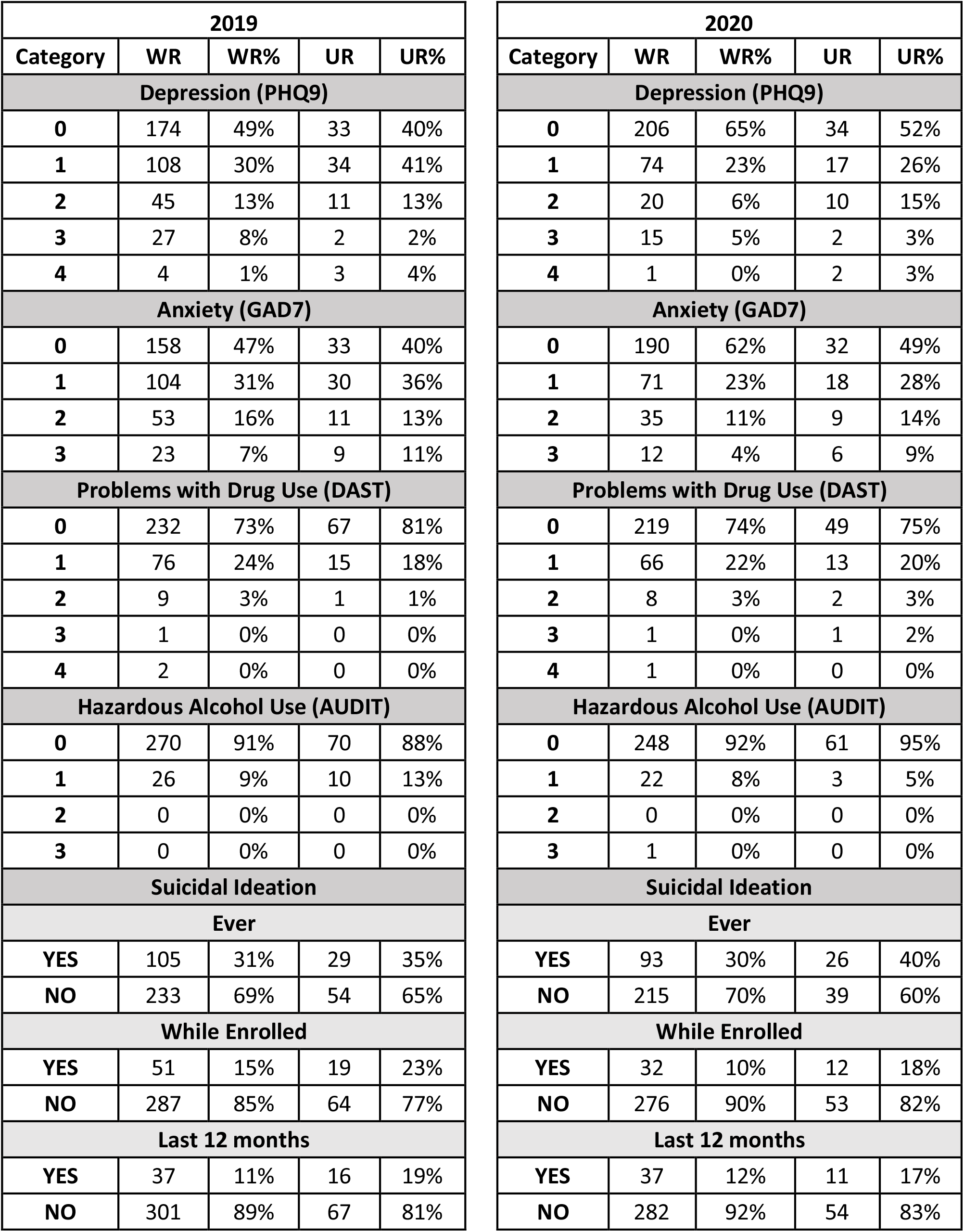
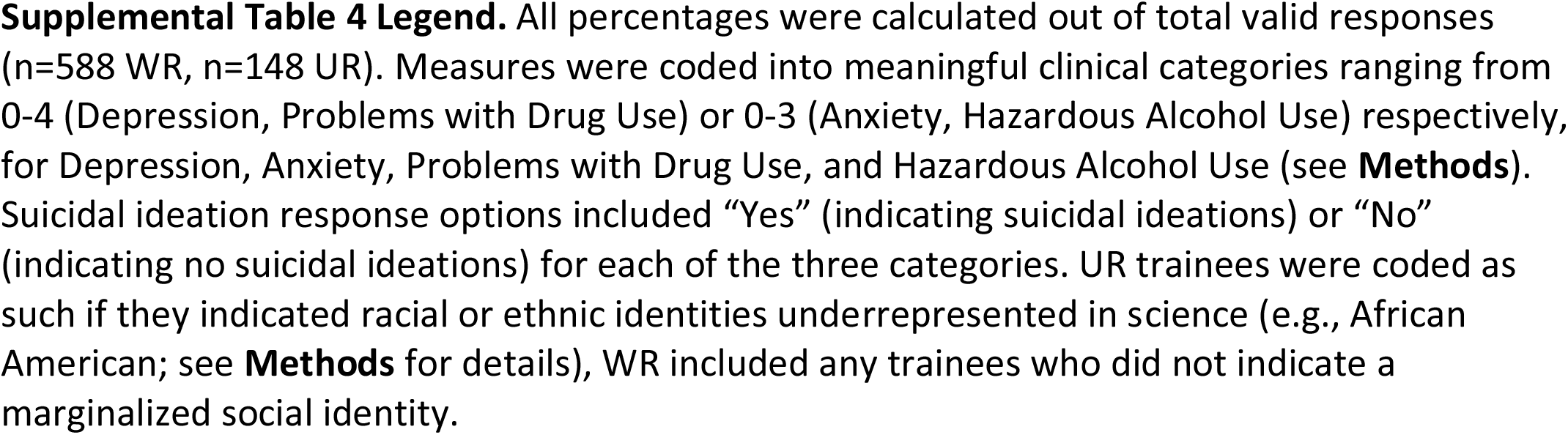
**Depression, Anxiety, Problems with Drug Use, Hazardous Alcohol Use, and Suicidal Ideation for WR vs. UR Trainees**

## Appendix 7. Supplemental Table 3a & 3b

**Supplemental Table 3a.**
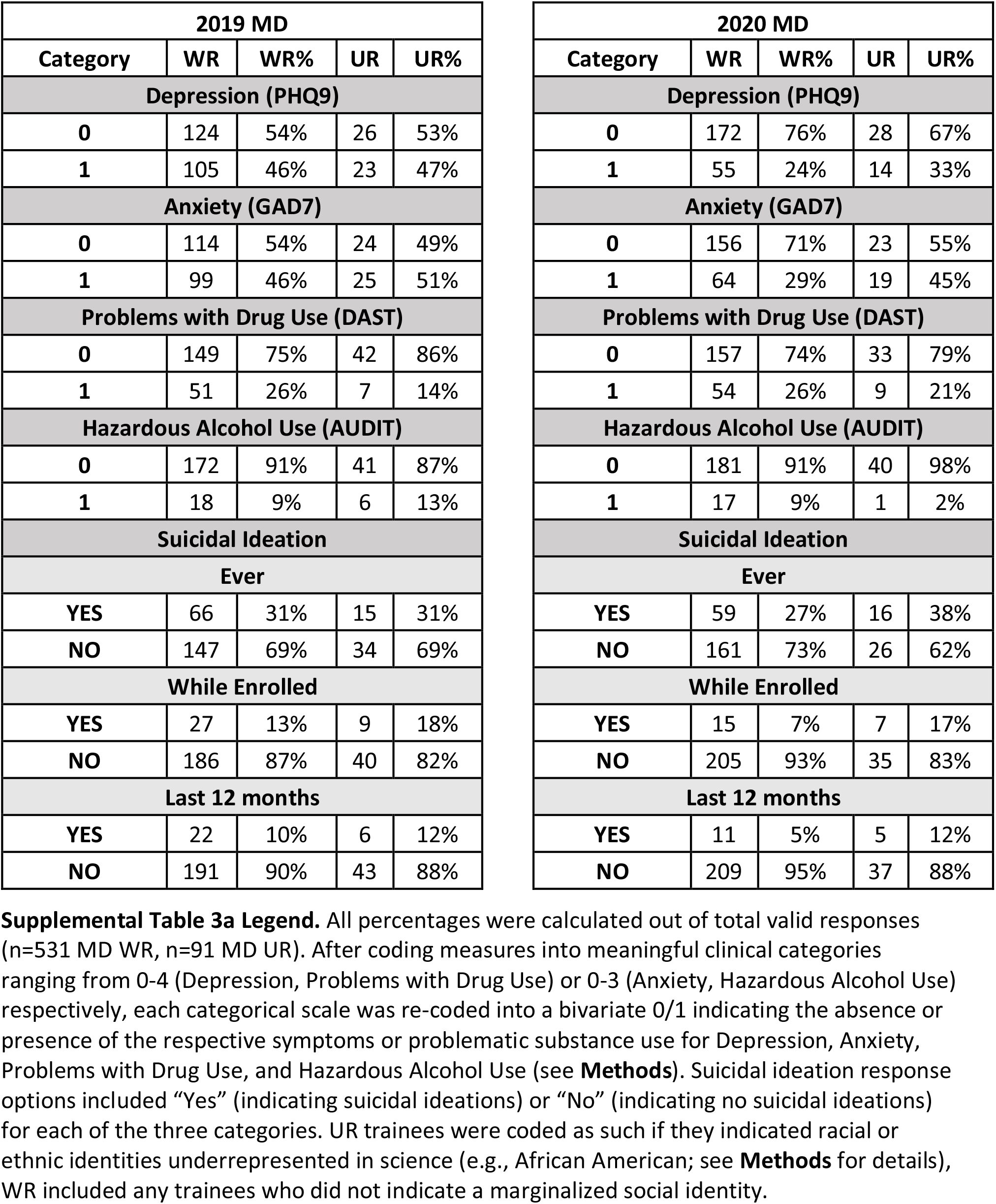
**Depression, Anxiety, Problems with Drug Use, Hazardous Alcohol Use, and Suicidal Ideation for MD WR vs. UR Trainees**

| 2019 MD |  |  |  |  |
| --- | --- | --- | --- | --- |
| Category | WR | WR% | UR | UR% |
| **Depression (PHQ9)** |  |  |  |  |
| 0 | 124 | 54% | 26 | 53% |
| 1 | 105 | 46% | 23 | 47% |
| **Anxiety (GAD7)** |  |  |  |  |
| 0 | 114 | 54% | 24 | 49% |
| 1 | 99 | 46% | 25 | 51% |
| **Problems with Drug Use (DAST)** |  |  |  |  |
| 0 | 149 | 75% | 42 | 86% |
| 1 | 51 | 26% | 7 | 14% |
| **Hazardous Alcohol Use (AUDIT)** |  |  |  |  |
| 0 | 172 | 91% | 41 | 87% |
| 1 | 18 | 9% | 6 | 13% |
| **Suicidal Ideation** |  |  |  |  |
| **Ever** |  |  |  |  |
| YES | 66 | 31% | 15 | 31% |
| NO | 147 | 69% | 34 | 69% |
| **While Enrolled** |  |  |  |  |
| YES | 27 | 13% | 9 | 18% |
| NO | 186 | 87% | 40 | 82% |
| **Last 12 months** |  |  |  |  |
| YES | 22 | 10% | 6 | 12% |
| NO | 191 | 90% | 43 | 88% |

| 2020 MD |  |  |  |  |
| --- | --- | --- | --- | --- |
| Category | WR | WR% | UR | UR% |
| **Depression (PHQ9)** |  |  |  |  |
| 0 | 172 | 76% | 28 | 67% |
| 1 | 55 | 24% | 14 | 33% |
| **Anxiety (GAD7)** |  |  |  |  |
| 0 | 156 | 71% | 23 | 55% |
| 1 | 64 | 29% | 19 | 45% |
| **Problems with Drug Use (DAST)** |  |  |  |  |
| 0 | 157 | 74% | 33 | 79% |
| 1 | 54 | 26% | 9 | 21% |
| **Hazardous Alcohol Use (AUDIT)** |  |  |  |  |
| 0 | 181 | 91% | 40 | 98% |
| 1 | 17 | 9% | 1 | 2% |
| **Suicidal Ideation** |  |  |  |  |
| **Ever** |  |  |  |  |
| YES | 59 | 27% | 16 | 38% |
| NO | 161 | 73% | 26 | 62% |
| **While Enrolled** |  |  |  |  |
| YES | 15 | 7% | 7 | 17% |
| NO | 205 | 93% | 35 | 83% |
| **Last 12 months** |  |  |  |  |
| YES | 11 | 5% | 5 | 12% |
| NO | 209 | 95% | 37 | 88% |

**Supplemental Table 3b.** **Depression, Anxiety, Problems with Drug Use, Hazardous Alcohol Use, and Suicidal Ideation for PHD WR vs. UR Trainees**

| 2019 PHD |  |  |  |  |
| --- | --- | --- | --- | --- |
| Category | WR | WR% | UR | UR% |
| Depression (PHQ9) |  |  |  |  |
| 0 | 50 | 39% | 7 | 21% |
| 1 | 79 | 61% | 27 | 79% |
| Anxiety (GAD7) |  |  |  |  |
| 0 | 44 | 35% | 9 | 26% |
| 1 | 81 | 65% | 25 | 74% |
| Problems with Drug Use (DAST) |  |  |  |  |
| 0 | 83 | 69% | 25 | 74% |
| 1 | 37 | 31% | 9 | 26% |
| Hazardous Alcohol Use (AUDIT) |  |  |  |  |
| 0 | 98 | 91% | 29 | 88% |
| 1 | 10 | 9% | 4 | 12% |
| Suicidal Ideation |  |  |  |  |
| Ever |  |  |  |  |
| YES | 39 | 31% | 14 | 41% |
| NO | 86 | 69% | 20 | 59% |
| While Enrolled |  |  |  |  |
| YES | 24 | 19% | 10 | 29% |
| NO | 101 | 81% | 24 | 71% |
| Last 12 months |  |  |  |  |
| YES | 15 | 12% | 10 | 29% |
| NO | 110 | 88% | 24 | 71% |

| 2020 PHD |  |  |  |  |
| --- | --- | --- | --- | --- |
| Category | WR | WR% | UR | UR% |
| Depression (PHQ9) |  |  |  |  |
| 0 | 34 | 38% | 6 | 26% |
| 1 | 55 | 62% | 17 | 74% |
| Anxiety (GAD7) |  |  |  |  |
| 0 | 34 | 39% | 9 | 39% |
| 1 | 54 | 61% | 14 | 61% |
| Problems with Drug Use (DAST) |  |  |  |  |
| 0 | 62 | 74% | 16 | 70% |
| 1 | 22 | 26% | 7 | 30% |
| Hazardous Alcohol Use (AUDIT) |  |  |  |  |
| 0 | 67 | 92% | 21 | 91% |
| 1 | 6 | 8% | 2 | 9% |
| Suicidal Ideation |  |  |  |  |
| Ever |  |  |  |  |
| YES | 34 | 39% | 10 | 43% |
| NO | 54 | 61% | 13 | 57% |
| While Enrolled |  |  |  |  |
| YES | 17 | 19% | 5 | 22% |
| NO | 71 | 81% | 18 | 78% |
| Last 12 months |  |  |  |  |
| YES | 15 | 17% | 6 | 26% |
| NO | 73 | 83% | 17 | 74% |

